# CoViD-19 in Italy: a mathematical model to analyze the epidemic containment strategy and the economic impacts

**DOI:** 10.1101/2020.05.28.20115790

**Authors:** Fabio Verachi, Luca Trussoni, Luciano Lanzi

## Abstract

The objective of this paper is to evaluate the potential costs deriving from the adoption of the CoViD-19 epidemic management strategy. For this purpose, we developed a specific methodology that combines an epidemiological model, known in the literature as “SIR” (Susceptible - Infected - Recovered), and a probabilistic state model, also known as “multi-state”. The model thus conceived was then parameterized using the dataset published by the Italian Government through the Civil Protection and the Istituto Superiore di Sanità. We therefore estimated the duration of the disease and the related costs, with reference to the strategy currently under discussion between government institutions and social organizations involved. Given the flexibility of the adopted approach, the tool will also be able to provide useful indications in relation to any alternative strategies that the Government could adopt in the near future, as well as being the starting point of an analysis of the epidemic indirect costs such as losses of GDP fractions.

## State of art

### CoViD19 epidemic and modeling works

As recently in the spotlight, the CoViD-19 epidemic refers to the so-called coronavirus disease, technically caused by SARS-CoV-2 virus (coronavirus 2 from severe acute respiratory syndrome), found to be similar to at least 70% of its gene sequence to that of SARS-CoV. Initially identified in the city of Wuhan in December 2019, the capital of the Chinese province of Hubei, it subsequently spreads to more than 210 countries in the world.^4^

The first confirmed cases of the virus in human patients were found towards the end of November 2019. From the middle of January 2020, the first cases were also found outside of China, brought by international travelers, mainly to the major trading partner nations of the country. The following table shows the spread of CoViD-19 for the main countries affected by the infection (as of April 19^th^, 2020).^5^

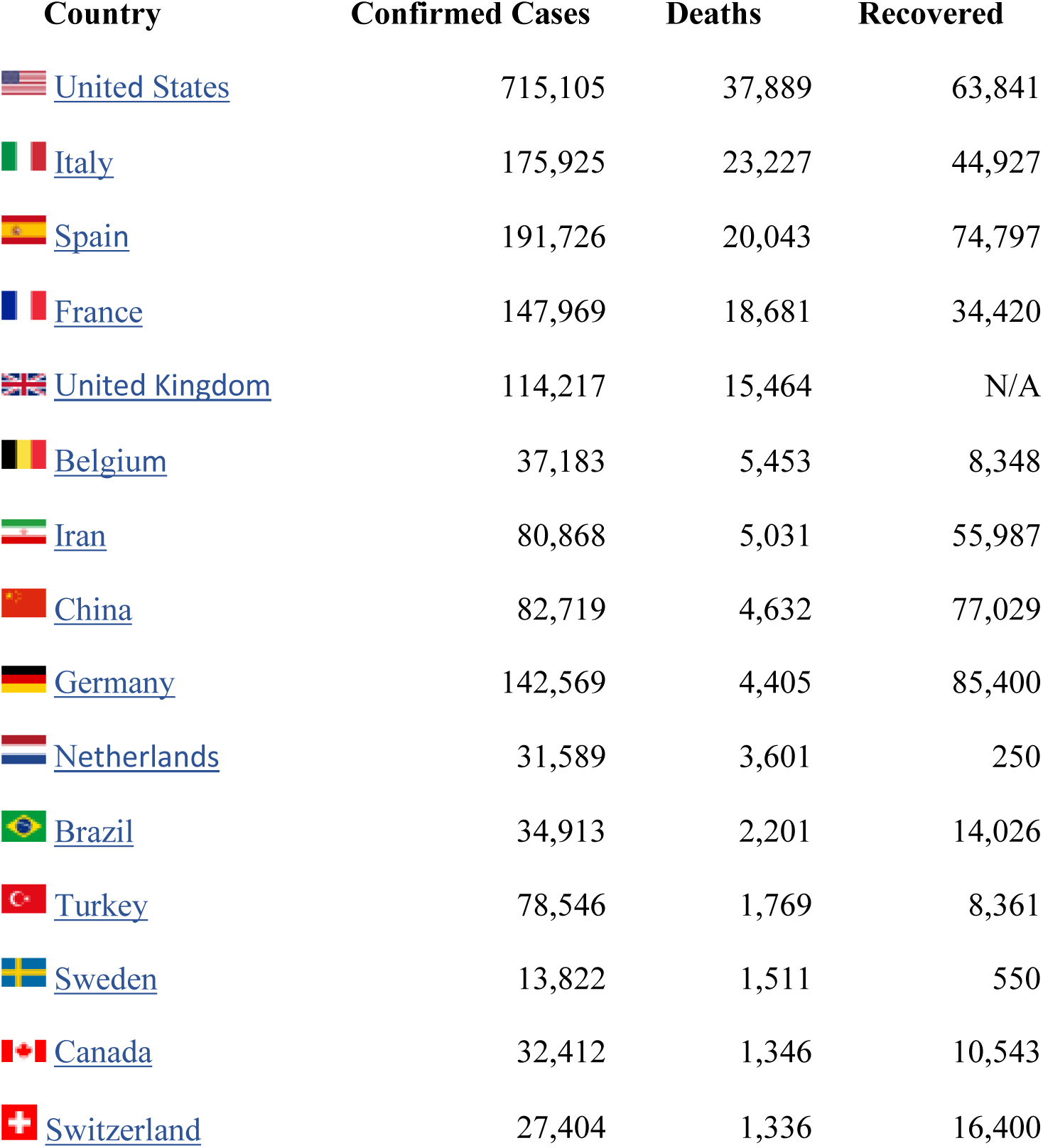

### SIR and SEIR models

The methodology proposed in this paper for the epidemiological component is a variant of the SIR (Susceptible, Infected, Recovered) model. The use of an epidemiological model is of fundamental importance to quantitatively define the infection dynamics.

It is also necessary to determine the possible strategies for identifying, preventing and managing the disease as well as, as will be seen, to estimate the direct and indirect economic impacts.

The SIR model was first proposed in the 1920s by Kermack and McKendrick to explain the rapid growth and subsequent decrease in the number of infected people observed in some epidemics such as the London plague in 1665-1666, Bombay in 1906 or London cholera in 1865.

The logic behind the methodology is to distinguish, among the population, the susceptible individuals (the uninfected but susceptible to infection) and the infected individuals (infected people who are able to transmit the disease).

The underlying mathematical model is a system of three differential equations:

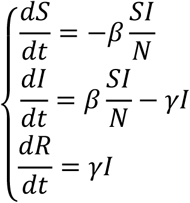

where *S* represents the population susceptible to the disease, *I* the infected population and *R* the “removed” population that is no longer able to spread the disease (composed by the healed and immunized people and the deceased people). The constants *β* and *γ* identify the evolution of the disease (respectively an infectivity parameter and the reciprocal of the average time of evolution of the disease).

Many variations have been introduced over time, such as the SEIR models, which explicitly consider the class of “exposed” people: this model is particularly valid for diseases with a significant incubation time, during which the infected are not yet contagious:

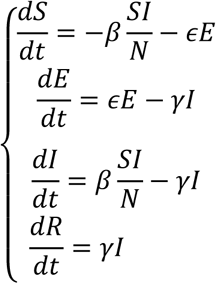

where the parameter *ϵ* represents the reciprocal of the incubation time. Further variants arise from the explicit consideration of time delays. For the CoViD-19 epidemic, researchers have proposed both the direct application of the SIR model (Nesteruk [22]) and the SEIR model (Zhou et al. [16]), variants of the SEIR with additional components (Tang et al. [11]), the explicit modeling of external factors (such as injection or subtraction of population due to travel Wu et al. [3]) or in which those exposed contribute to the spread of the disease ([16]).

For the sake of completeness, alternative modeling approaches not based on differential equations should be mentioned, among them the stochastic models, typically based on discrete-time agent models (the differential models are continuous-time): these approaches will not be used in this paper.

### A proposal for an integrated model

The model we propose consists of two sub-models that can be independently defined: an epidemiological model (of the SIR type, briefly described above), and a state model that describes the management of the infected people. The SIR model estimates the speed of spread and the size of the epidemic (in line with Batista, [15] and [21]), while the state model is a proposal from the authors, analogous to models used in the credit risk assessment, through which we will arrive at an estimate of the epidemic management needs (in terms of hospitalization costs).

The choice of a SIR-type model is motivated both by the literature findings and by the fact that there is a broad consensus that even during the incubation period, people exposed to SARS-Cov2 virus should be considered infectious.

### Structure of the SIR model

The model is characterized by the following dynamics:

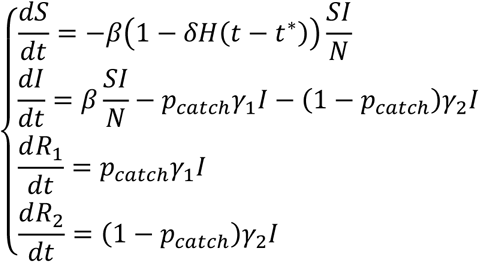

with the following meaning of the variables:

- *N* represents the consistency of the population in which the epidemic develops;
- *S* represents, in every moment of time, the “susceptible” population, which has not been infected by the disease;
- *I* represents the population that has been infected with the disease and that is active in the spreading phenomenon;
- *P_catch_* represents the probability that a case will be caught by the disease management system;
- *γ*_1_ and *γ*_2_ respectively represent the reciprocals of the average times during which the infected spread the disease. In the model, it will be assumed 1/*γ*_1_ = 5.2 days and 1/*γ*_2_ = 15 days (in line with the indications in Li et al. [2]);
- *R*_1_ and *R*_2_ represent the population of the “removed”, or those who are no longer affected by the epidemic (because they are isolated, healed or deceased), respectively if they have been intercepted or not by the disease management system;
- *β* represents the speed of free spread of the epidemic;
- *δ* represents the effectiveness of the lockdown in limiting the spread of the epidemic, starting from the time *t^*^* corresponding to March 8^th^, 2020. The function *H* is the classic Heaviside function (*H*(*x*) = 1 se *x ≥* 0, *H*(*x*) = 0 se *x* < 0).

A classic parameter of epidemiological models is the “basic reproduction number” that represents the average number of susceptible individuals converted into infected for each individual infected, usually indicated with the acronym *r*_0_.

The proposed model is characterized by three *r*_0_ parameters, one for the “catched” population *r*_0_*_,catched_ = β/γ*_1_, one for the “non-catched” population *r*_0,_*_non catched_ = β/γ*_2_, and an overall estimation *r*_0_ *= β/*(*p_catch_γ*_1_ + (1 − *p_catch_*)*γ*_2_).

The method followed to adapt the model to the observed data is described in the Appendix, and leads to identification of the following values:

| Parameter | Value | Parameter | Value |
| --- | --- | --- | --- |
| $N$ | 1,995,998 | $\delta$ | 60.0% |
| $\beta$ | 0.2770 | $t^*$ | 38 |
| | | $p_{catch}$ | 5.38% |

In the values above we can implicitly infer the “basic reproductive numbers”, equal to *r*_0_ = 4.01*; r*_0,_*_catched_ =* 1.44*; r*_0_*,_non catched_ =* 4.16 before the lockdown and *r*_0_ = 2.41*; r*_0_*,_catched_ =* 0.86*; r*_0_*_,non catched_ =* 2.50 after it. The values, although high, do not disagree with the literature which in particular reports, for the pre-lockdown values, different ranges referring to different geographical areas^6^:

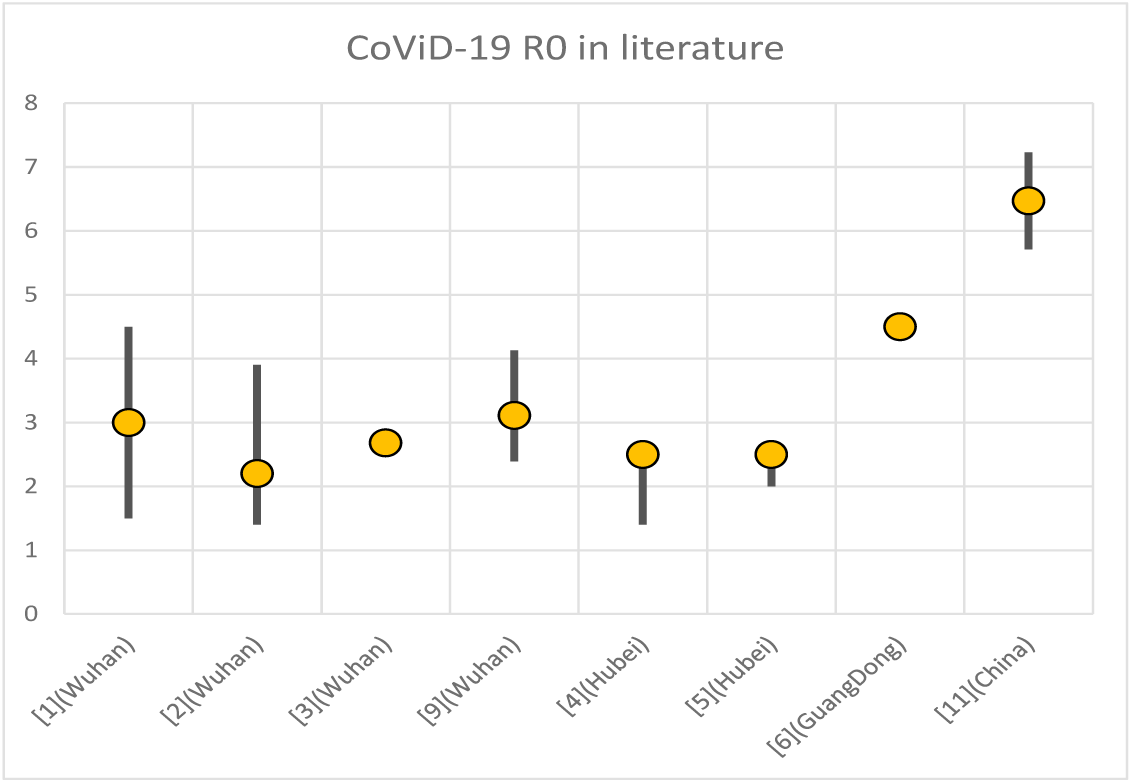

Below we report some charts to compare the results of the model with observations:

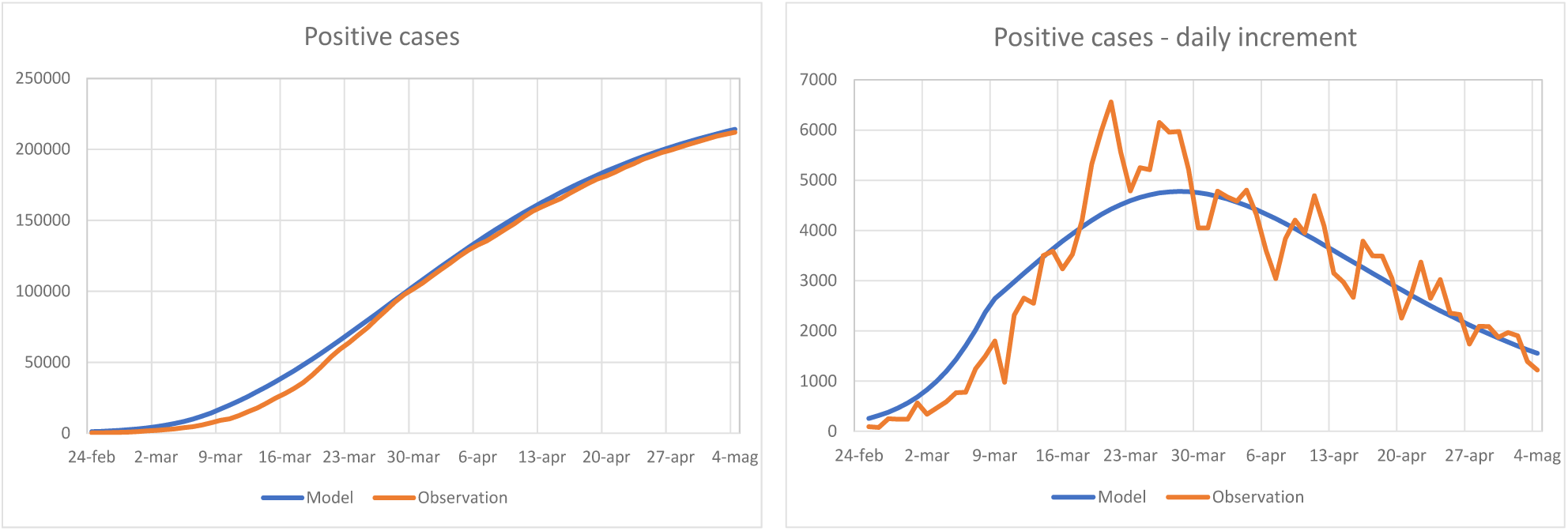

We point out that the model seems to support the conclusion that the lockdown, however effective in reducing the *r*_0_, was not enough to reduce it below value one in not-catched cases. This phenomenon can be explained by the fact that the main consequence of the lockdown is to have locked the value of *N* at two million people, instead of allowing it to rise up to the entire Italian population of 60 million people.

It follows that the circulation of the disease remains high within the “confined” but not isolated population. This conclusion seems plausible to the authors and in line with other studies: an extreme case was the Diamond Princess cruise ship, where the illness circulation exhibited an *r*_0_ equal to about 15 (Rocklov et al. [7]).

The authors consider a high level of *r*_0_ reasonable for the spread of the disease within the confined population, also in light of the preliminary study of the Istituto Superiore di Sanità which highlights how the contagions in the period 1^st^ - 23^rd^ April 2020 in Italy are for 2/3 occurred in rest homes and in the family environment (ISS [26]).

This population must be considered as the “circle” of the infected, within which the lockdown was only partially effective (think of the case of those people infected in home isolation who certainly no longer infect their work colleagues but expose their family members to a significant risk of contagion).

The value of *p_catch_* is also remarkable, the interpretation of which is that the model implies the presence of about 18.6 not symptomatic/not recognized infection cases for each case intercepted by the national health service. Different estimates have been proposed for asymptomatic to symptomatic infection rate ratio: for example, a 5:1 ratio for the Wuhan province has been proposed by Day [24], while Li et al. [25] propose arange between 5:1 and 10:1. On the other hand, Wu et al. [27] find a 94% asymptomatic cases for the Chinese outbreak, in line with our result.

Finally, the value *t^*^* implies that the model estimates the beginning of the circulation of the disease in Italy on January 17^th^ (initiation level at 10 infected).

### Patient management model

The patient management model focuses on the evolution of the *R*_1_(*t*) aggregate of the SIR model. The basic idea stems from the observation that during the accumulation phase, visually stable relationships were observed between the deaths and the number of patients in intensive care on the previous day.

Such empirical observations suggested that patient management could be modeled using approaches known in the literature as “multi-state”. A multi-state model describes how a single individual moves between a series of states in discrete time. Movements in a space of discrete states is governed by the transition intensity matrix which represents the instantaneous risk of moving from one state to another state.

In our application this means that the transition of the health status of the subjects takes place through successive states of illness depending on its duration.

The following model was therefore formulated:

- for each head that enters in the aggregate *R*_1_ at time *t* three periods are identified *M*_1_ = [*t, t + T*_1_], *M*_2_ = [*t + T*_1_*,t + T*_2_], *M*_3_ = [*t* + *T*_2_, + ∞];
- at any instant of simulated time the head can be in one of five states (*R*_1_*_G_, R*_1_*_D_, R*_1_*_Hm_, R*_1*HS*_, *R*_1_*_ICU_*), which respectively represent the patients recovered, deceased, in home isolation, hospitalized or in intensive care;
- for each period of illness *M_i_* we define the probabilities 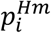, 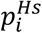, 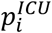 that the head (not healed or deceased) is in one of the three states on any day of the period;
- for each of the periods *M_i_* is defined a differentiated probability of daily death *p_D,Mi_;* the heads pass to the deceased state according to the length of the period (in particular, in each period the deaths are calculated by applying a first in first out logic);
- for the periods *M*_2_ and *M_3_* two daily probabilities of recovery *p_H, Mi_* are specifically defined; the heads pass to the healed state according to the length of the period (again utilizing the same first in first out logic).

The model is therefore identified by a total of 13 parameters (in the appendix the structure of the model and the technique used for parameter fitting are detailed).

With regard to the parameters, the following values are obtained:

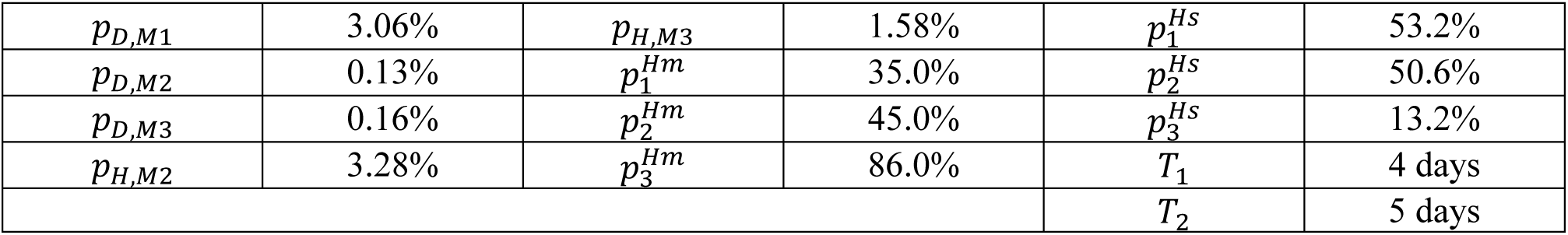

From parameters, complementing to 1, we can easily deduce the probabilities of being treated in intensive care in the three periods 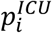, respectively 11.8%, 4.4% and 0.8%.

The model exhibits a good fit with the observed values, as highlighted by the following charts (we called Hospital ratio the fraction of hospitalized infected compared to the intercepted infected, and ICU ratio the fraction of hospitalized patients treated in intensive care):

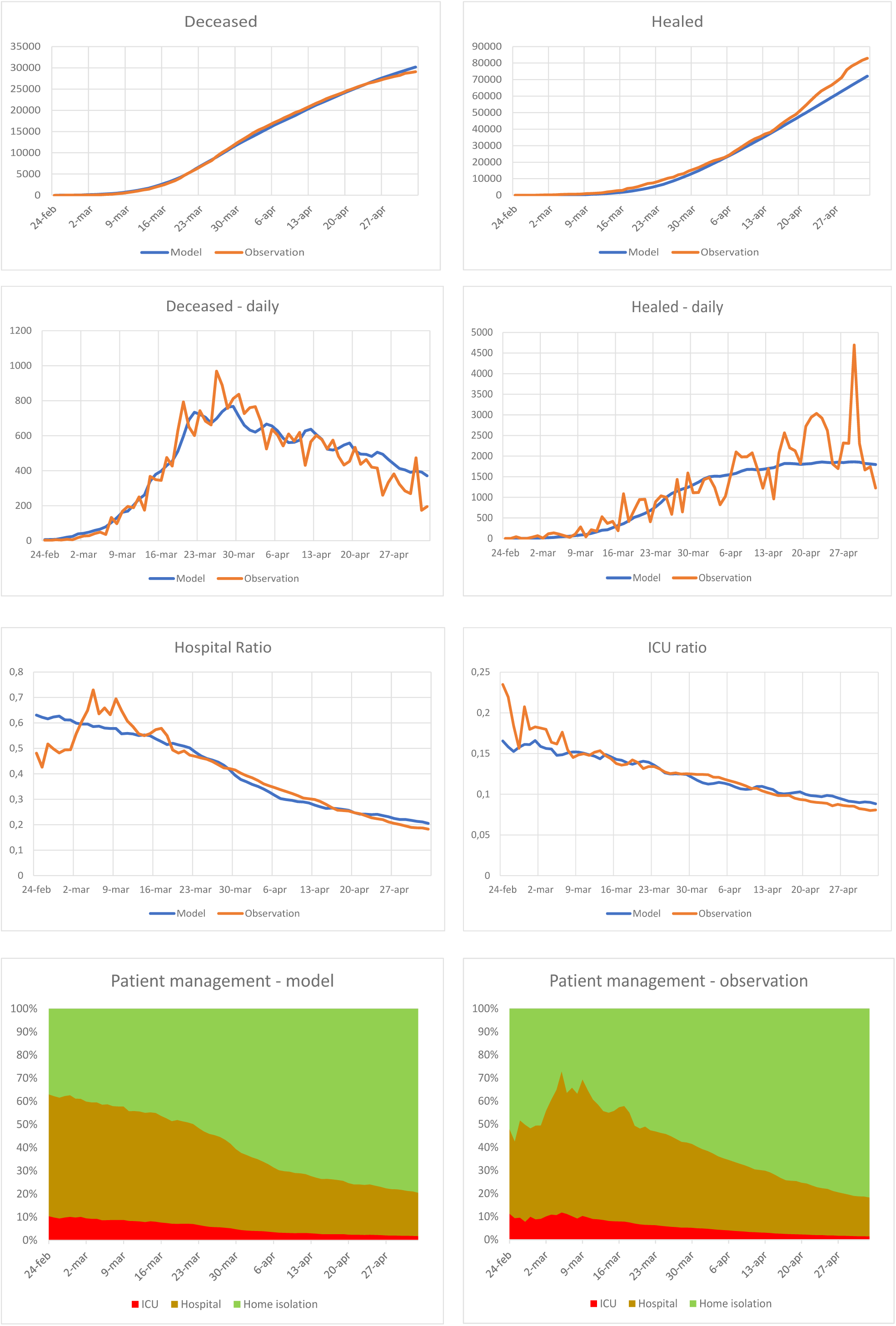

With regard to the parameters, we point out that the following relationships should hold:

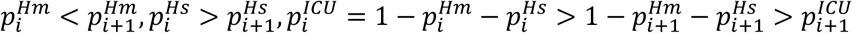

All these relations are consistent with the reasonable hypothesis that patients who survive for a greater number of days have a progressively more favorable prognosis.

The satisfactory agreement with the observed data and further tests carried out by the authors on trends at regional level, using models with the same structure, depose in favor of the reliability of the proposed approach: however, all the usual limitations and cautions in using mathematical models remain applicable in representing reality.

A substantial improvement in the analysis of the model’s performance would occur using the comparison with the data relating to the actually observed clinical evolution of the patients: these data are not available while writing this paper.

### Integration of models and simulations

For the purpose of fitting, the patient management model is fed with the observed increments: to integrate the models it is sufficient to feed the model with the simulated increases of the *R*_1_ population taken from the SIR model.

In the following sections, all the simulations are carried out on a total of 430 simulated days.

First of all, we note that the simulated evolution of the epidemic takes place essentially in the first 200 simulated days:

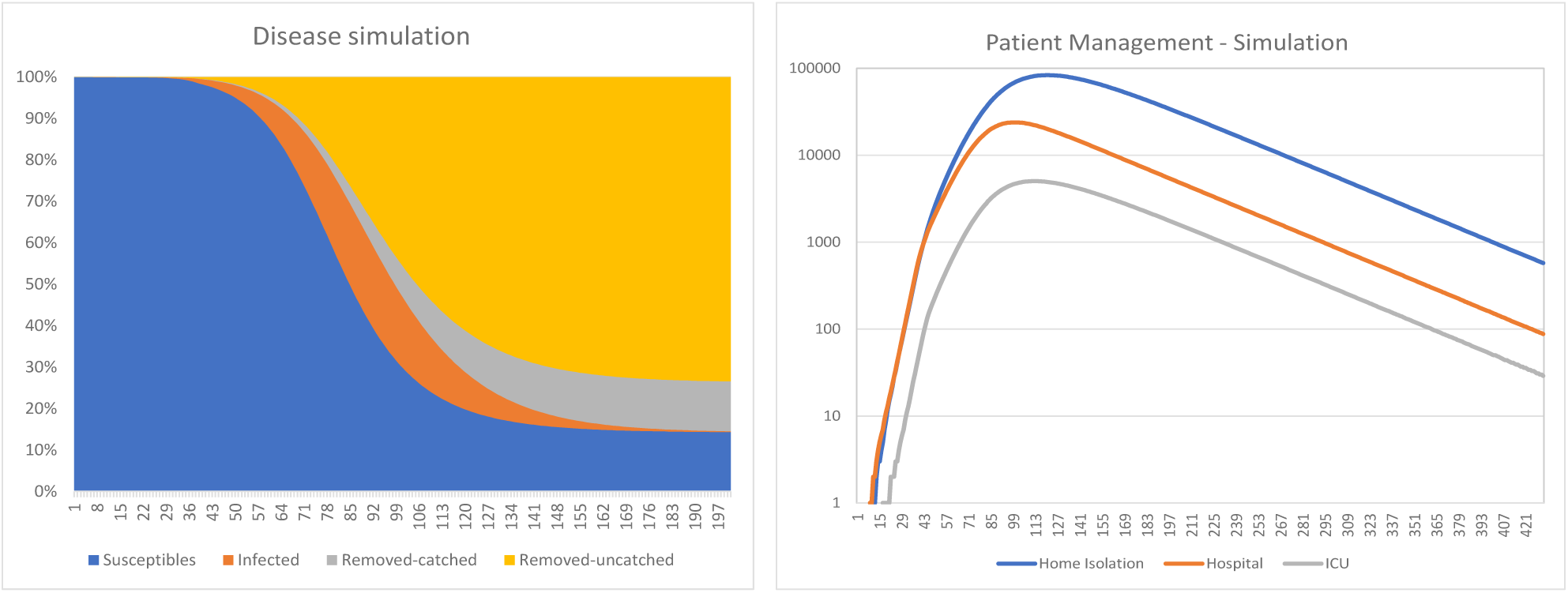

By applying the patient management model, the peaks in the number of patients in ICU, hospital and home isolation happen at day 116, 102 and 123 respectively (with a timespan, from the first to the last peak, of 21 days). As of the writing date of this article, the maximum of the classes of hospitalized patients have already been observed (Aprl 3^th^ and 4^th^), while the maximum of the infected in home isolation has been observed on May 2^nd^ with and observed delay of 29 days from the first peak.

It is also possible to calculate some relevant quantities, such as the number of deaths, the number of days in the various management classes, as well as the duration of the epidemic until reaching predetermined levels of the infected population (remember that infected people spread the disease).

The following values are obtained:

|  | 430 days (simulation) | As at May 4<sup>th</sup> | % |
| --- | --- | --- | --- |
| Total deaths | 46,011 | 29,079 | 63% |
| ICU days occupied | 563,497 | 157,238 | 28% |
| Hospital days occupied | 2,325,604 | 1,232,943 | 53% |
| Home isolation days occupied | 9,676,022 | 2,978,830 | 31% |
| Time to 10,000 infected |  | 181 | days |
| Time to 1,000 infected |  | 228 | days |
| Time to 100 infected |  | 274 | days |
| Time to 10 infected |  | 320 | days |

The previous table also shows the progress compared to May 4^th^ (which would be the 108th simulated day). Assuming a standard cost of 500 euros for a day of normal hospitalization and 1,500 euros for a day of hospitalization in intensive care, the direct costs of the epidemic should be around 2 billion euros.

A useful exercise is then to apply the model to different sets of initialization parameters, varying the “size” of the population *N* and the delay in activating the restrictive measures.

The following values are obtained:

| Dimension | Delay | Deaths | Hospital days | ICU days | Isolation days | Duration to |  |  |  |
| --- | --- | --- | --- | --- | --- | --- | --- | --- | --- |
|  |  |  |  |  |  | 10000 infected | 1000 infected | 100 infected | 10 infected |
| 10.000 | 15 | 198 | 2.988 | 12.307 | 52.724 |  | 86 | 137 | 183 |
| 100.000 | 15 | 2.267 | 28.303 | 116.672 | 485.957 | 111 | 162 | 209 | 256 |
| 1.000.000 | 15 | 23.004 | 281.531 | 1.162.254 | 4.833.101 | 188 | 234 | 280 | 327 |
| 5.000.000 | 15 | 114.997 | 1.405.601 | 5.805.454 | 24.126.683 | 237 | 283 | 331 | 377 |
| 10.000 | 30 | 208 | 3.094 | 12.728 | 54.556 |  | 71 | 114 | 157 |
| 100.000 | 30 | 2.279 | 28.449 | 117.275 | 488.585 | 93 | 143 | 189 | 235 |
| 1.000.000 | 30 | 23.034 | 282.056 | 1.164.088 | 4.843.180 | 168 | 215 | 263 | 309 |
| 5.000.000 | 30 | 115.084 | 1.407.892 | 5.812.470 | 24.171.071 | 220 | 265 | 313 | 359 |
| 10.000 | 45 | 227 | 3.290 | 13.523 | 57.965 |  | 67 | 103 | 139 |
| 100.000 | 45 | 2.391 | 29.877 | 123.203 | 513.544 | 80 | 121 | 162 | 202 |
| 1.000.000 | 45 | 23.281 | 285.298 | 1.177.216 | 4.899.544 | 148 | 194 | 239 | 285 |
| 5.000.000 | 45 | 115.431 | 1.412.816 | 5.830.999 | 24.259.517 | 200 | 246 | 292 | 339 |

The underlying idea of the exercise is to simulate the reopening of an epidemic event in our country (Italy), due to the emergence of an outbreak during a hypothetical phase of release of the restrictive measures. Assuming as the duration of the lockdown the time from the delayed detection of the outbreak to the return to the threshold of 1,000 infected people, the following relationship could be empirically found:

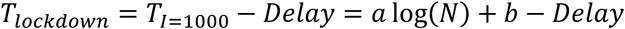

with *a =* 30.64 and *b = −*208.82. It is reasonable to assume the existence of an exponential relationship between the infected population size *N* and the lockdown delay^7^: observing that *N* of the adopted model is equal to about 2 million people reached with a detection delay of 38 days, we can find the following relation:>

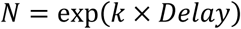

where *k =* 0.38. By replacing this formula in the expression for the duration of the lockdown, we are able to find that each day of delay in the detection costs about 11 days of lengthening the lockdown and that, neglecting the effects of saturation, a detection delay of 47 days would be sufficient to expand the infected people until the total national population amount.

## Conclusions

This paper focused on defining and applying a model for the management of the CoViD-19 epidemic. The phenomenon has been studied based on the Italian experience, with the purpose of identifying a preliminary estimate for the economic impacts of the disease. As previously described, the methodology we propose consists of two different independent models to be used jointly: the epidemic dynamics model (SIR) and a multi-state model for patient management.

The first model (SIR) seems to intercept pretty well the dynamics that characterize the contagion, especially in terms of basic reproduction number (*r*_0_) and the number of people in whom the epidemic develops (*N*). As highlighted from the empirical evidence, captured by the model, it appears that the basic reproductive number of the epidemic *r*_0_ for uncatched cases remains above 1 even during the lockdown. The authors believe that this phenomenon is essentially linked to the interaction between the fitting procedure and the nature of the lockdown: on one side the model is free to identify the dimension of the population within the disease spreads, and obviously concentrates on the fraction of population within active diffusion is detected, on the other the lockdown limited the size of the potentially exposed population through which the disease spread but was not effective in slowing down the diffusion within the confined population clusters (best known cases are the outbreaks in retirement homes RSA throughout Italy). It should also be noted that, in the model considered, asymptomatic infected patients, whose number seems to be high compared to symptomatic cases, took a relevant role in the spread dynamics.

The second model (multi-state) focused instead on the relationships that take into account the transition of patients through the possible duration of the different stages of the disease, represented respectively by patients recovered, deceased, in home isolation, hospitalized or in intensive care. By considering this information together with the estimates of the SIR model, it is possible to obtain a forecast of the duration in terms of days of lockdown to reach a residual threshold (e.g. 100 infected patients).

We also proposed a scenario analysis related to the spread of the disease in smaller confined populations, and a preliminary cost measure of the “delay” of activation of the lockdown measures in terms of the duration of the lockdown itself: these analyzes, resulting from an extrapolation of the calibrated model (and therefore subject to the limits of precision and reliability of the same), could constitute a basis for the modeling of future outbreaks that may reoccur during the subsequent phase of relaxation of the current lockdown constraints.

However, an estimate of the direct costs of the epidemic is a partial, and in some ways secondary, aspect of the total economic impact: the loss of Italy’s GDP fractions caused by the lockdown, to be assessed with the necessary sector specificities, is to be considered the main effect of the epidemic. The authors believe that the proposed model, allowing to obtain indications on the duration of the lockdown, could also be useful for estimating indirect impacts, to which we will eventually dedicate future analyzes.

## Data Availability

The fitting of the epidemiological model is obtained starting from the data made available by the Civil Protection through the GitHub repository (https://github.com/pcm-dpc/COVID-19/blob/master/dati-andamento-nazionale/dpc-covid19-ita-andamento-nazionale.csv.) that covers, as of the writing date of this article, the period from February 24th to May 4th 2020

## Appendix – Technical details on the models

### Fitting of the epidemiological model

The fitting of the epidemiological model is obtained starting from the data made available by the Civil Protection through the GitHub repository [23] that covers, as of the writing date of this article, the period from February 24^th^ to May 4^th^ 2020^8^. A first consideration is that the SIR model considered has a diffusion parameter for the pre-lockdown period and a different parameter for the post-lockdown period: the post-lockdown period, however, being made up of dates after March 8^th^ 2020 included, has a data intensity much higher than the previous one (40 points versus 13), and therefore the estimates made for this second period are to be considered more stable. The SIR parameter estimation process is made of the following steps:

I. we define an auxiliary SIR model based on the equations *dS/dt = — β^*^SI/N; dI/dt = +β^*^SI/N − p_catch_γ*_1_*I −* (1− *p_catch_*)*γ*_2_*I; dR*_1_*dt = p_catch_γ*_1_*I*; modeling of component *R*2 at this level is ignored; the analysis compares the model with historical data for the lockdown period, or for data subsequent to the instant *t^*^* assumed equal to March 8^th^. Furthermore, the parameters *γ*_1_ and *γ*_2_ are locked at 5.2 days and 15 days (in line with the literature indications for the time to onset of symptoms and for the evolution of the disease in shorter cases; time to onset of symptoms is used in the first case since the fraction of the population routed to the first class of the removed is intercepted and isolated at the onset of symptoms, while in the second case the infected remain infectious and asymptomatic for the entire duration of the disease), the initial state is: *S*(0) = *S*_0_; *I*(0) = *I*_0_; *R*_1_(0) = 7,535, where 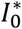 and 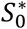 are both identified by the model;
II. we fit the SIR model based on equations *dS/dt = −β*(1 − *δH*(*t − t^*^*))(*SI/N*) *; dI/dt = + β*(1 − *δH*(*t − t^*^*))(*SI/N*) − *p_catch_γ*_1_*I* − (1 − *p_catch_*)*γ*_2_*I; dR*_1_*/dt = p_catch_γ*_1_*I; dR*_2_*/dt* = (1 − *p_catch_*)*γ*_1_*I*. Both the parameters *γ*_1_ and *γ*_2_ are locked as in the previous step, and the further constraint *βδ = β^*^* is introduced in order to benefit from the greater amount of information of the second period. The parameter *t^*^* is also left free so as to allow the model to infer the initial instant of evolution of the disease, conventionally placed at *S*(0) *= N −* 10,*I*(0) = 10, *R*_1_(0) = *R*_2_(0) = 0.

In both cases the dynamic used is a gradient descent. The fitting parameters of step I are used as the starting point for step II.

The fitting of both steps is performed through the minimization of the *L*_1_ norm, calculated on the total number of cases detected versus the time series of the simulated population *R*_1_(*t*). Defined 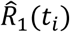 the time series of the cases observed by the Civil Protection, the fitting procedure seeks the minimum value of the norm 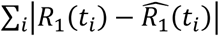. A commonly used approach is the maximization of the log-likelihood of the increases: it is assumed that the quantities Δ*R*_1_(*t_i_*) *= R*_1_(*t_i+_*_1_) − *R*_1_(*t_i_*) represent the expected values *λi* of a random Poisson variable whose corresponding observation is 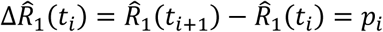 and then the fitting procedure look for the minimum of the norm 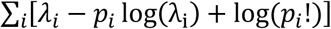. In principle, different combinations of the two approaches are possible by performing steps I and II for one norm or the other:

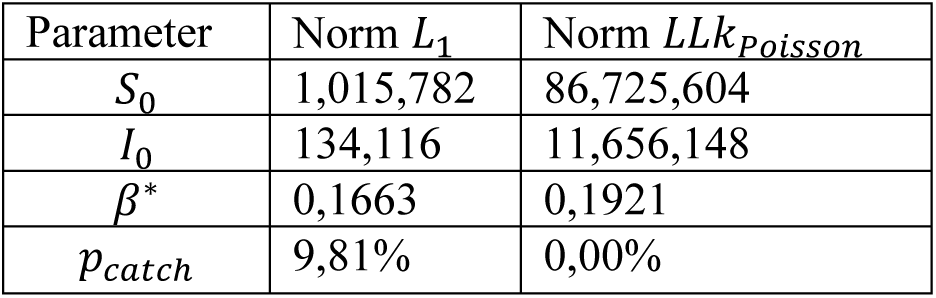

Using the obtained parameters as:

| Parameter | I. Norm $L_1$<br>II. Norm $L_1$ | I. Norm $L_1$<br>II. Norm $LLk_{Poisson}$ | I. Norm $LLk_{Poisson}$<br>II. Norm $L_1$ | I. Norm $LLk_{Poisson}$<br>II. Norm $LLk_{Poisson}$ |
| --- | --- | --- | --- | --- |
| $t^*$ | 38 | 51 | 33 | 20 |
| $N$ | 1,995,898 | 60,000,000<br>(active limit) | 24,802,511 | 56,694,585 |
| $\delta$ | 60,0% | 68,0% | 47,5% | 39,6% |
| $p_{catch}$ | 5,38% | 0,15% | 0,38% | 0,15% |

The authors opted for the strategy Norm *L*_1_+ Norm *L*_1_ both for consistency between the two steps and for the reasonability on the dimension of the population in which the disease spread, that, looking at the death toll, should not be in the tens of millions range.

### Fitting the multi-state model

For the state model, the strategy is to minimize the norm *L*_1_:

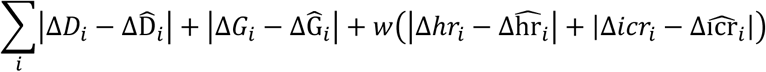

where Δ*D_t_* and 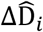 respectively represent the expected and observed daily deaths, the values Δ*G_i_* and 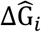 the expected and observed daily healings, the values Δ*hr_i_* and 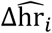; the hospital ratio expected and observed, Δ*icr_i_* and 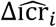 the planned and observed intensive care ratio, and *w* a weight that has the purpose of connecting the scales of the variables (assumed equal to 500). The hospital ratio is defined as the ratio between the patients hospitalized against the total number of infected people (for each moment of time) and the intensive care ratio is defined as the ratio between the patients admitted to intensive care against the patients hospitalized (both at internal and outside of intensive care). The model is powered by the observed inputs 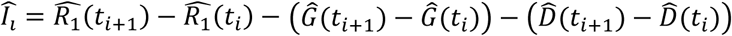 and initialized by dividing the 229 cases detected at February 24^th^ into 5 classes (4 of 46 cases and one of 45 cases) distributed over the first 5 days of population development. The norm is calculated on the 71 increments observed from February 24^th^ to May 4^th^, 2020.

### Multi-state model implementation code

This section shows the pseudocode that implements the multi-state model: the objective is an operational description of the model, which allows its possible replication. It is assumed that the outputs of the SIR epidemiological model are accumulated in the vectors `R1[t], R2[t]`. The population state at every moment of time is accumulated in the array of vectors `ActiveCatched[t] [i]`, where the vectors `ActiveCatched[t]` are supposed to be of variable length.

~~~
ActiveCatched[1]=[0]
For any t in 2…length(R1[t])
     EnteringR=R1[t]-R1[t-1]
     ActiveCatched[2..N+1]=ActiveCatched[1..N]
     AcriveCatched[1]=EnteringR
     Base1=sum(ActiveCatched[t-1][1…T1])
     Base2 = sum(ActiveCatched[t-1][T1…T2])
     Base3=sum(ActiveCatched[t-1][T2…])
     ActiveCatched[t]=FIFOOut(ActiveCatched[t], (*p*_D,_*_M_*_3_*+p_H,M_*_3_) *Base3)
     ActiveCatched[t]=FIFOOut(ActiveCatched[t], (*p*_D_,_M2_+*p*_H,M2_) **Base2,T2)
     ActiveCatched[t]=FIFOOut(ActiveCatched[t], *p_D,M_*_1_ *Base1,T1)
     DeathEvents[t] = (Base1*p_D,M1_+Base2\**p*_D_,_M2_+Base3\**p*_D_,_M3_)
     HealEvents[t]= (Base2\**p_H,M_*_2_*+*Base3*\*p_HM_*_3_)
~~~

The FIFO out function eliminates people from the active case vector by subtracting them from those of older age:

~~~
FIFOOut(vector,events,index)
     If index not specified
          Index=length(vector)
     Aux=events
     For i=index to 1
          If vector[i]<aux
               Aux=aux-vector[i]
               Vector[i]=0
          Else
               Vector[i]=vector[i]
               Aux=0
     If Aux>0 throw exception(“Too many events!”)
~~~

The execution of the previous code segment defines the vectors `DeathEvents` and `HealEvents`, representing deaths and daily healings respectively. To obtain the daily consistency of the populations in home isolation, hospitalized or in intensive care, the following pseudocode is performed:

~~~
for any t in 1…length(ActiveCatched)
     Base1=sum(ActiveCatched(1…T1))
     Base2 = sum(ActiveCatched(T1…T2))
     Base3=sum(ActiveCatched(T2…))
     PopulationVector=[Base1,Base2,Base3]
     HomeIsolated[t]=PopulationVector•Home_V
     Hospital[t]=PopulationVector•Hosp_V
     ICU[t]=PopulationVector•ICU_V
~~~

where 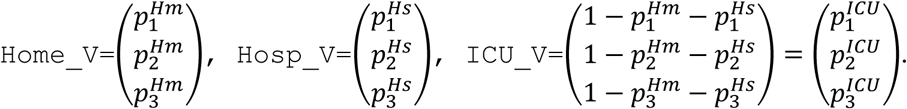

`HomeIsolated, Hospital` and `ICU` are vectors that contain, at any time, the population subjected to home isolation, hospitalization and intensive care treatment respectively.

## Footnotes

4 Further details in David S. Hui, Esam EI Azhar, Tariq A. Madani, Francine Ntoumi, Richard Kock, Osman Dar, Giuseppe Ippolito, Timothy D. Mchugh, Ziad A. Memish, Christian Drosten e Alimuddin Zumla “The continuing 2019-nCoV epidemic threat of novel coronaviruses to global health – The latest 2019 novel coronavirus outbreak in Wuhan, China” in International Journal of Infectious Diseases, vol. 91, 14 January 2020, pp. 264–266.

5 For confirmed cases, deaths and recovered in each country see “Confirmed Cases and Deaths by Country, Territory, or Conveyance” in Worldometer.

6 For example, in the Diamond Princess the *r*_0_ of the epidemic spread is estimated at 14.8 [7]. The authors wish to cite theGabriel Goh epidemic simulator, https://gabgoh.github.io/COVID/index.html, in whose documentation can be found the values that have been used for the reported graph.

7 The dynamic is consistent with the hypothesis that each simulated person has a constant number of contacts with new people for each simulated day. This hypothesis is reasonable in estimating a population only as long as saturation effects are not achieved.

8 An early version of this paper appeared on AIFIRM website, https://www.aifirm.it/news/da-leggere/, in which models were fitted using data from February 24^th^ to April 16^th^.

